# Patient, Socioeconomic, and Clinical Influences on Clinical Trial Participation in Stargardt Disease

**DOI:** 10.1101/2025.08.24.25334317

**Authors:** Dorothy T Wang, Bani Antonio-Aguirre, Annabelle Pan, Maria Ludovica Ruggeri, Setu P Mehta, Christy H Smith, Kelsey S Guthrie, Carolyn Applegate, Jefferson J Doyle, Mandeep S Singh

## Abstract

**Importance:** Multiple clinical trials for Stargardt disease are ongoing, as there is no treatment available for this condition. However, timely recruitment of clinical trial participants remains challenging, risking incomplete study cohorts and prolonged study timelines that together escalate the possibility of trial failure. Factors associated with clinical trial participation in Stargardt are poorly understood. Identifying drivers of clinical trial participation and genetic testing completion may improve trial design and access.

**Objective:** To identify demographic and clinical factors associated with clinical trial participation and genetic testing completion in Stargardt.

**Design:** Retrospective cohort study of patients seen at a genetic eye disease center from January 2003 to December 2024. Associations were evaluated using bivariate analysis and multivariable logistic regression.

**Setting:** Single-center study at a tertiary referral center.

**Participants:** 280 patients clinically diagnosed with Stargardt disease [median (IQR) age at presentation 37 (20-52) years; follow-up 4.5 (1.6-10.1) years].

**Exposures:** Demographic and clinical factors, including race, insurance, education, employment, parenting status, age of onset, and genetic counseling history.

**Main Outcomes and Measures:** Genetic testing completion (all participants) and clinical trial participation (among *ABCA4-*positive participants).

**Results:** Among 280 participants, 246 (88%) completed genetic testing. Among 223 *ABCA4*-positive participants, 45 (20%) had enrolled in a clinical trial. Clinical trial participation was associated with later onset [≥60 years; OR (95% CI) 36.7 (2.5-537.1)], part-time employment [9.3 (1.01-172.9)], and was less likely among retired individuals [0.014 (0.0005-0.4)] or those without genetic counseling [0.22 (0.07-0.71)]. In multivariable analysis, genetic testing completion was less likely in participants who were Black [0.40 (0.16-0.98)], under/uninsured [0.47 (0.19-0.97)], without genetic counseling [0.09 (0.03-0.22)], or not planning to have children [0.35 (0.14-0.89)]. Common reasons for non-completion included loss to follow-up, refusal, and awaiting genetic counseling.

**Conclusions and Relevance:** In this large single-center cohort, one in five participants with molecularly confirmed Stargardt had enrolled in a clinical trial. Older age of onset, part-time employment, and genetic counseling may promote clinical trial participation among Stargardt patients, while retirement may be negatively associated. These findings may ultimately support the development of rational strategies to accelerate clinical trial enrollment, thereby broadly supporting the development of effective treatments for Stargardt disease.

## Introduction

Stargardt disease (STGD) is the most common form of hereditary macular dystrophy, with an estimated global prevalence of 1:6562.^1^ It is primarily caused by autosomal recessive mutations in the *ABCA4* gene, leading to toxic bisretinoid accumulation in retinal pigment epithelium cells and progressive bilateral central vision loss that typically begins in the first or second decade of life.^2–4^

Genetic testing (GT) plays a vital role in diagnosing and managing inherited retinal diseases (IRDs) such as STGD. GT provides diagnostic confirmation, prognosis estimation, inheritance pattern identification, screening of at-risk family members, and eligibility determination for interventional trials. Additionally, GT will be crucial for determining eligibility for gene-specific therapies as they become available.^5^ Despite these benefits, access to GT has been historically limited by factors such as high costs, insurance restrictions, and variability in provider familiarity with testing protocols.^5–7^

In the United States (U.S.), no-cost GT initiatives, including eyeGENE and the Foundation Fighting Blindness My Retina Tracker GT Study (FFB-MRT-GTS), have expanded access for IRD patients.^8–10^ However, significant barriers remain, particularly among underrepresented populations. Disparities in referral patterns and access to genetic counseling (GC) may contribute to unequal access to GT and, consequently, to investigational therapies.^6^ While there are a few studies on GT patterns in IRDs considered as a broad disease class, those specifically examining STGD remain scarce.^9,11^ To our knowledge, no large-scale U.S. study has evaluated demographic and clinical factors associated with GT completion specifically in STGD populations.

In parallel, numerous therapeutic strategies for STGD—including gene therapy, stem cell therapy, optogenetics, and pharmaceuticals— are under investigation, though no FDA-approved treatments currently exist.^12,13^ As such, clinical trial (CT) participation represents the only pathway for many patients to access investigational therapies. However, efficient recruitment of study participants remains a challenge, thus impeding rapid study completion. Understanding the factors associated with CT participation among people with STGD may help in implementing rational strategies to increase the efficiency of CT participant recruitment.

This study aims to address these gaps by evaluating the demographic and clinical factors associated with GT completion and CT participation in a large U.S. cohort of patients with STGD.

## Methods

### Ethics

This study was conducted at the Wilmer Eye Institute, Johns Hopkins Hospital, with institutional review board (IRB) approval (IRB00213488). It adhered to the tenets of the Declaration of Helsinki and complied with Health Insurance Portability and Accountability Act regulations. Informed consent for this retrospective medical record review was not required by the Johns Hopkins University IRB.

### Data collection and patient selection

We conducted a retrospective cohort study of patients seen at the Wilmer Eye Institute’s Genetic Eye Disease (GEDi) center between January 2003 and December 2024. Patients were included if they had a clinical diagnosis of STGD confirmed by a fellowship-trained IRD specialist. Individuals with nonspecific diagnoses of retinal, macular, or cone-rod dystrophies without documented suspicion of STGD were excluded.

Demographic variables were extracted from the electronic medical record and included sex, race/ethnicity, insurance type, employment status, education level, English proficiency, and parenting status. Race and ethnicity were categorized using National Institutes of Health (NIH) guidelines as American Indian or Alaska Native, Asian, Black or African American, Hispanic or Latino, Native Hawaiian or Other Pacific Islander, White, or Other.^14^ Insurance status was collected as private, public, international self-pay, or uninsured; for regression analyses, this was dichotomized into private vs. under/uninsured (including public, international self-pay, and uninsured), reflecting differing access and coverage in the U.S. healthcare system.

Clinical variables included age of symptom onset, age at presentation, duration of follow-up, family history of STGD or other IRDs, history of consanguinity, and baseline and follow-up best-corrected visual acuity (BCVA). BCVA was recorded in Snellen and subsequently converted to logarithm of the minimum angle of resolution (logMAR) for analysis.^15^ The better-seeing eye was defined as the eye with lower logMAR, and the worse-seeing eye as the eye with higher logMAR. If both eyes had identical BCVA, the same value was assigned to both categories. Participants who had completed genetic testing prior to baseline evaluation from our institution were excluded from all analyses involving baseline BCVA measurements.

GT status was recorded as binary (yes/no). For untested participants, reasons for non-completion were categorized as: awaiting genetic counseling, awaiting IRD retinal specialist evaluation, lost to follow-up, participant declined testing, testing not covered by insurance, or other.

### CT participation

CT participation was assessed among participants with positive or likely positive GT results in *ABCA4* (ABCA4-GT). Positive ABCA4-GT was defined as two pathogenic *ABCA4* variants, either confirmed *in trans* or of unknown phase. Likely positive ABCA4-GT included cases with one pathogenic variant plus one VUS; two VUS/novel variants *in trans*; or one pathogenic variant (or two *in cis*) with a typical STGD phenotype, per the published ProgStar criteria.^16^ Classification was based on manual review of clinical laboratory reports by a trained grader.

Comprehensive chart review identified CT participation through clinical notes, imaging/procedure reports, and the media section using keyword searches (e.g. “clinical trial,” “NCT,” “study/screening visit,” “study/screening protocol”) and review of informed consent documents. CT participation was recorded as a binary variable, and trial-specific enrollment was extracted. Identified trials included both Johns Hopkins (JH)-based and external studies: TEASE (NCT04239625; non-JH), ProgSTAR/SMART (NCT01977846; JH), SeaSTAR (NCT03772665; JH), POLARIS (NCT06435000; JH), oral metformin (NCT04545736; non-JH), Zimura (NCT03364153; JH), docosahexaenoic acid (DHA) supplementation (NCT00060749; non-JH), and Stargazer (NCT04489511; non-JH).

### Statistical analysis

De-identified data were analyzed using Microsoft Excel (version 16.84.24041420; Microsoft Corp) and Stata v.18.0 (Stata Corporation LLC, College Station, USA). Missing data resulted from the absence of relevant documentation in medical records and led to variable denominators. For descriptive and bivariate analyses, participants were excluded only from analyses of variables with missing data. Multivariable logistic regression used complete case analysis, excluding participants with missing data for any covariate included in the model.

Bivariate analyses compared participants with and without GT completion. The Shapiro-Wilk test assessed normality of continuous variables; Mann-Whitney U was used for non-normally distributed continuous variables. Chi-square or Fisher’s exact tests were used for categorical comparisons, with Fisher’s applied when expected counts were <5. Multivariable logistic regression models assessed predictors of GT completion (full cohort) and CT participation (*ABCA4*-positive subgroup), with variable selection via least absolute shrinkage and selection operator (LASSO) regression and reference groups selected by the highest frequency for categorical variables. Statistical significance was set at α = 0.05.

## Results

### Demographic characteristics of Stargardt participants

Among 280 STGD participants, 46% were male, and the median age was 47 years (interquartile range [IQR] 30.5-63; range 4-88). Most identified as White (62%), followed by Black or African American (26%), Asian (10%), American Indian or Alaska Native (2.5%), and Native Hawaiian or Other Pacific Islander (<0.5%); 4% reported Other race. Hispanic or Latino ethnicity was reported by 4%.

Overall, 86% had or planned to have children; 14% reported neither having children nor plans to have children. Insurance status included 73% with private, 21% public, 5% international self-pay, and 1% uninsured. Employment data (n=247) showed 52% employed full-time, 19% students, 15% retired, 6% each on disability or unemployed, and 2% employed part-time.

Among 213 with education data, 57% held a university degree, 15% had postgraduate education, 13% had a high school diploma or General Educational Development (GED), 11% were children enrolled in age-appropriate schooling, 3% held trade school or associate degrees, and 1% completed only primary school. For analysis, education was grouped as post-secondary (84%) vs. primary or secondary (16%); children enrolled in age-appropriate schooling were excluded to minimize potential confounding from parental educational background and decision-making. English was used in 96% of clinic visits.

Bivariate analysis showed significant differences in GT completion by race (23% of tested vs. 47% of untested participants identified as Black; p=0.002), insurance type (p=0.047), education (p=0.021), and parenting status (p=5E-4) (**Table 1)**.

**Table 1.** Demographic characteristics of participants clinically diagnosed with Stargardt disease (N=280), stratified by genetic testing status. Data presented as % (n); denominators vary due to missing data. Statistical significance was assessed using chi-squared tests (χ^2^) for variables with expected counts ≥5 and Fisher’s exact tests (two-tailed p-values) for those <5. ^a^Participants identifying as multi-racial may be counted in more than one race category. ^b^“Under/Underinsured” includes public insurance, no insurance, or international self-pay. ^c^Post-secondary education includes university, associate/trade school, or postgraduate education. Children at age-appropriate education levels were excluded from this analysis due to potential confounding by parents’ highest education levels. Abbreviations: *PI* Pacific Islander

| Parameter | Total (N=280) | Tested cohort (N=246) | Untested cohort (N=34) | P-values (tested vs. untested) |
| --- | --- | --- | --- | --- |
| **Sex** | | | | $\chi^2$ p=0.350 |
| Male | 46% (128 of 280) | 47% (115 of 246) | 38% (13 of 34) |  |
| Female | 54% (152 of 280) | 53% (131 of 246) | 62% (21 of 34) |  |
| **Race<sup>a</sup>** |  |  |  |  |
| White | 62% (173 of 280) | 64% (157 of 246) | 47% (16 of 34) | $\chi^2$ p=0.059 |
| Black or African American | 26% (72 of 280) | 23% (56 of 246) | 47% (16 of 34) | $\chi^2$ p=0.002 |
| Native Hawaiian/Other PI | <1% (1 of 280) | <1% (1 of 246) | — | Fisher p=1.000 |
| Asian | 10% (28 of 280) | 11% (26 of 246) | 6% (2 of 34) | Fisher p=0.549 |
| American Indian/Alaska Native | 3% (7 of 280) | 3% (7 of 246) | — | Fisher p=1.000 |
| Other | 4% (12 of 280) | 4% (11 of 246) | 3% (1 of 34) | Fisher p=1.000 |
| **Ethnicity** |  |  |  | Fisher p=1.000 |
| Hispanic or Latino | 4% (11 of 280) | 4% (10 of 246) | 3% (1 of 34) |  |
| Not Hispanic or Latino | 96% (269 of 280) | 96% (236 of 246) | 97% (33 of 34) |  |
| **Type of insurance** |  |  |  | Fisher p=0.047 |
| Public insurance | 21% (59 of 280) | 19% (47 of 246) | 35% (12 of 34) |  |
| Private insurance | 73% (205 of 280) | 76% (186 of 246) | 56% (19 of 34) |  |
| International self-pay | 5% (13 of 280) | 4% (11 of 246) | 6% (2 of 34) |  |
| Uninsured | 1% (3 of 280) | 1% (2 of 246) | 3% (1 of 34) |  |
| **Insurance type (grouped)<sup>b</sup>** | | | | $\chi^2$ p=0.015 |
| Private insurance | 73% (205 of 280) | 76% (186 of 246) | 56% (19 of 34) |  |
| Under/Uninsured | 27% (5 of 280) | 24% (60 of 246) | 44% (15 of 34) |  |
| **Level of employment** |  |  |  | Fisher p=0.118 |
| Employed full-time | 52% (129 of 247) | 52% (116 of 221) | 50% (13 of 26) |  |
| Employed part-time | 2% (5 of 247) | 1% (3 of 221) | 8% (2 of 26) |  |
| Student | 19% (47 of 247) | 20% (44 of 221) | 12% (3 of 26) |  |
| Retired | 15% (37 of 247) | 14% (32 of 221) | 19% (5 of 26) |  |
| On disability | 6% (15 of 247) | 5% (12 of 221) | 12% (3 of 26) |  |
| Unemployed | 6% (14 of 247) | 6% (14 of 221) | — |  |
| **Highest level of education** |  |  |  | Fisher p=0.135 |
| Primary school | 1% (3 of 213) | 1% (2 of 191) | 5% (1 of 22) |  |
| High school diploma/GED | 13% (27 of 213) | 11% (21 of 191) | 27% (6 of 22) |  |
| Associate degree/trade school | 3% (6 of 213) | 3% (5 of 191) | 5% (1 of 22) |  |
| University degree | 57% (122 of 213) | 59% (112 of 191) | 45% (10 of 22) |  |
| Postgraduate education | 15% (32 of 213) | 16% (30 of 191) | 9% (2 of 22) |  |
| Child in appropriate schooling | 11% (23 of 213) | 11% (21 of 191) | 9% (2 of 22) |  |
| **Education level (grouped)<sup>c</sup>** |  |  |  | Fisher p=0.021 |
| Primary or secondary school | 16% (30 of 190) | 14% (23 of 170) | 35% (7 of 20) |  |
| Post-secondary school | 84% (160 of 190) | 86% (147 of 170) | 65% (13 of 20) |  |
| **English-speaking?** |  |  |  | Fisher p=1.000 |
| Yes | 96% (270 of 280) | 96% (237 of 246) | 97% (33 of 34) |  |
| No; requires clinical interpreter | 4% (10 of 280) | 4% (9 of 246) | 3% (1 of 34) |  |
| **Parenting status** |  |  |  | Fisher p=5E-4 |
| Have/may plan to have children | 86% (242 of 280) | 89% (220 of 246) | 65% (22 of 34) |  |
| No plans to have children | 14% (38 of 280) | 11% (26 of 246) | 35% (12 of 34) |  |

### Clinical characteristics and genetic testing (GT) history

Median age at symptom onset was 23 years (IQR 10-40), with presentation to GEDi at 37 years (IQR 20-52) and follow-up of 4.5 years (IQR 1.6-10.1). Family history of STGD or other IRDs was present in 41%; 3% reported consanguinity.

There were 38 participants who completed genetic testing prior to baseline evaluation at our institution; these participants were excluded from subsequent analyses with baseline BCVA. Median baseline BCVA was 0.602 logMAR (Snellen equivalent: 20/80) in the better-seeing eye and 0.796 (20/125) in the worse-seeing eye, worsening to 0.903 (20/160) and 1.00 (20/200), respectively, at follow-up. Participants who completed GT had slightly better BCVA at both timepoints, though not statistically significant (all p>0.06; see **Table 2**).

**Table 2.**
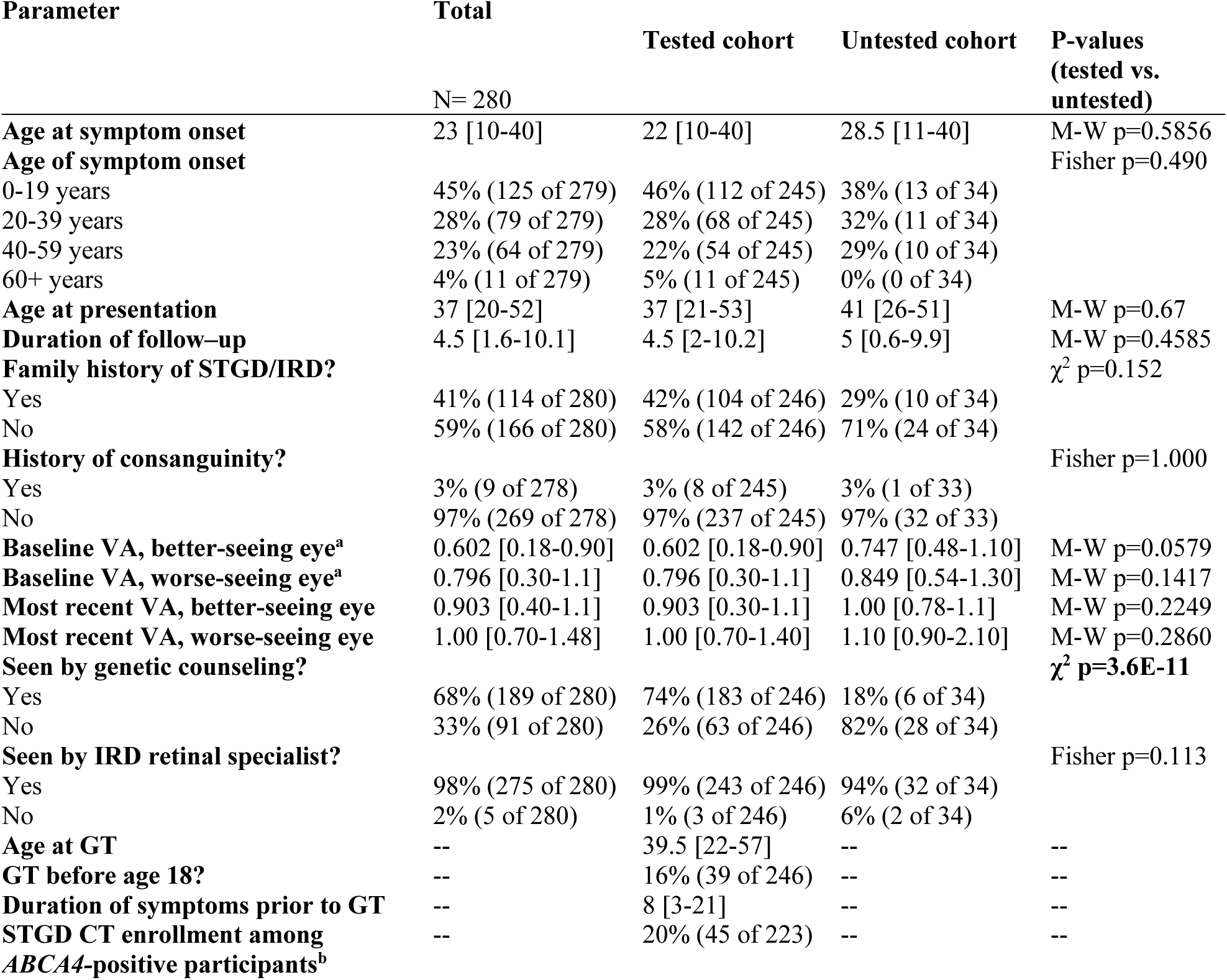
Clinical and genetic history characteristics of Stargardt disease participants (N=280), stratified by genetic testing status. Data are shown as % (n) or median [IQR]. Denominators vary due to missing data. All ages and durations are in years; BCVA values are in logMAR. As quantitative variables were non-normally distributed (Shapiro-Wilk test), comparisons used the Mann-Whitney U test. Categorical comparisons used chi-squared (χ^2^) or Fisher’s exact tests (two-tailed p-values), as appropriate. ^a^For baseline BCVA measurements (n=242), 38 participants who completed genetic testing prior to their baseline evaluation at our institution were excluded. ^b^*ABCA4-*positivity (n=223) was defined as positive or likely positive ABCA4*-*GT (see Methods for criteria) Abbreviations: *GT* genetic testing; *STGD* Stargardt; *IRD* inherited retinal disease; *IQR* interquartile range; *BCVA* best-corrected visual acuity (measured in logMAR); *M-W* Mann-Whitney U-test; *ABCA4-GT* ABCA4-specific genetic testing, refers to a manual reinterpretation of ABCA4-specific results using current variant databases and classification guidelines; *CT* clinical trial

Most participants (98%) were evaluated by an IRD specialist; 67.5% had seen a clinical genetic counselor (CGC). GT completion was significantly associated with GC (74% tested vs. 18% untested; p=3.6E-11).

Of the 246 participants who completed GT, the median age at GT was 39.5 years (IQR 22-57), with symptoms present for a median of 8 years (IQR 3-21) before testing. Sixteen percent underwent GT before age 18 (**Table 2**).

### Multivariable analysis of factors associated with genetic testing (GT) completion

In logistic regression (**Figure 1**; complete numerical values available in **eTable 1**), Black participants had significantly lower odds of GT completion than White participants (odds ratio [OR] 395; 95% confidence interval [CI] 0.160-0.876; p=0.044). Under/uninsured participants had lower odds than those with private insurance (OR 0.468; 95% CI 0.193-0.972; p=0.043). Those without children nor plans to have children had 65% lower odds of completing GT (OR 0.348; 95% CI 0.137-0.885; p=0.027) relative to those who had children or endorsed plans to have children. Lack of prior GC was strongly associated with lower odds of GT completion (OR 0.085; 95% CI 0.033-0.221; p=4.18E-7).

**Figure 1.**
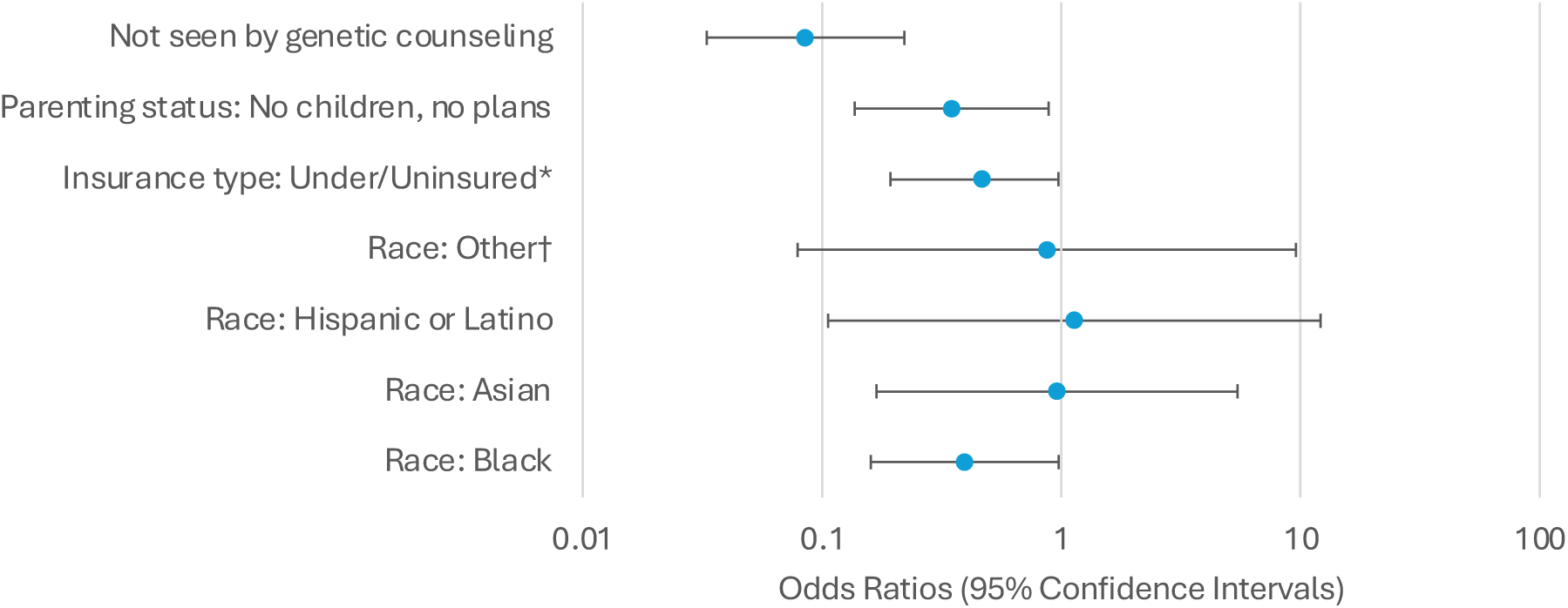
Multivariable analysis of factors associated with completing genetic testing among Stargardt disease participants (N=280) Least absolute shrinkage and selection operator (LASSO) regression was applied to select covariates for a binary logistic regression model. Odds ratios (ORs) and 95% confidence intervals (CIs) are plotted on a logarithmic scale, with a null value of OR = 1. *“Under/Uninsured” includes public insurance, no insurance, or international self-pay. †“Other” race was a composite category created for this analysis and includes Native Hawaiian or Other Pacific Islander and American Indian or Alaska Native, due to small sample sizes in each group. Reference groups were selected by highest frequency: Parenting status = have or may have children; Insurance type = private insurance; Race = White.

### Reasons for not completing genetic testing (GT) among untested participants

Among 34 untested participants, reasons for non-completion included loss to follow-up (41%), refusal (26%), awaiting genetic counseling (24%), awaiting retinal specialist evaluation (3%), and lack of insurance coverage for testing (3%) (**Figure 2**). One participant (3%), categorized as “Other,” cited difficulty performing home-based testing and expressed that GT was not a priority due to personal circumstances and the absence of children who might inherit the condition.

**Figure 2.**
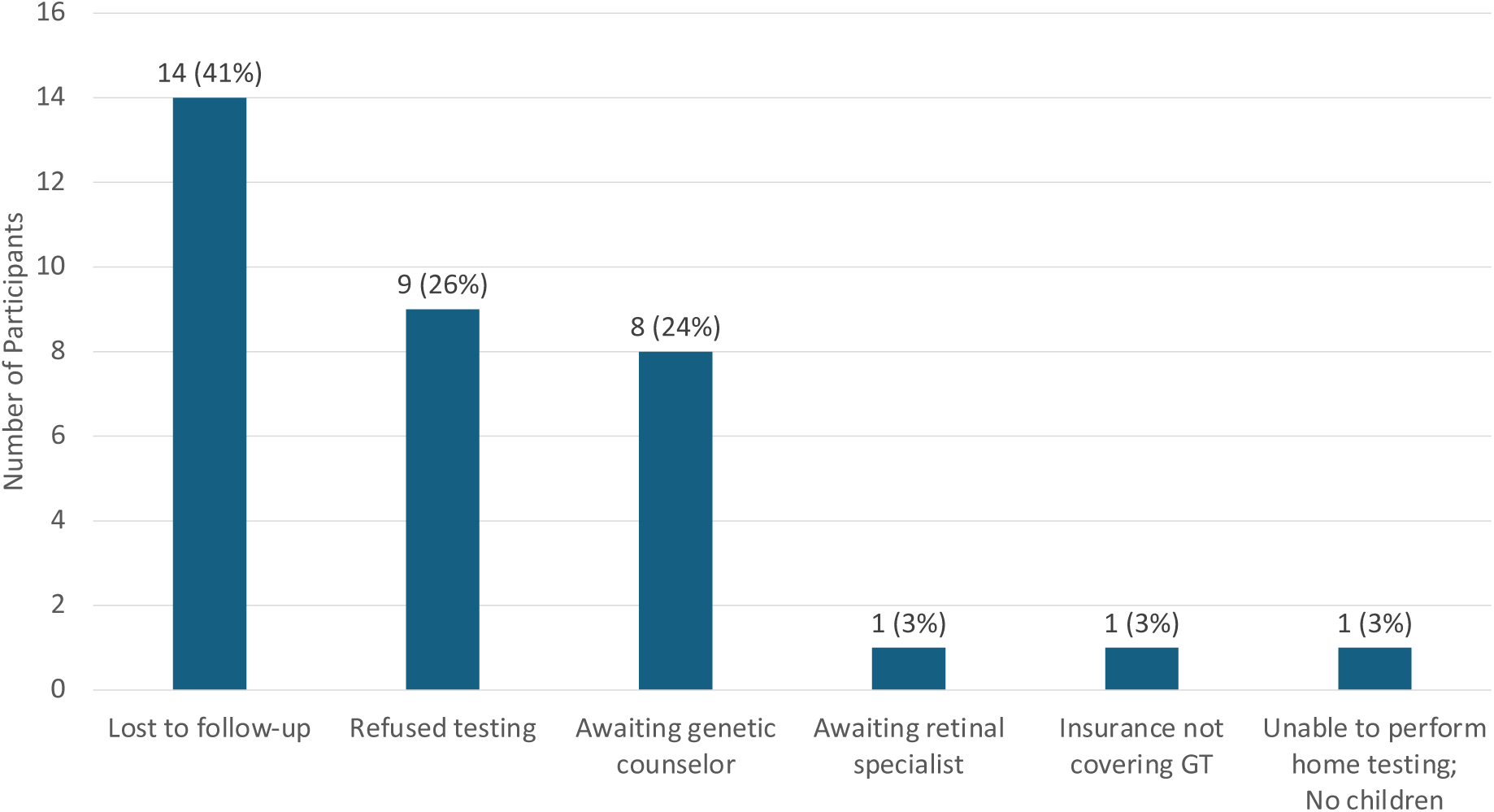
Reasons for not completing genetic testing among untested participants (N=34). Bars represent the number of participants categorized under each reason for not completing genetic testing (GT). Labels above each bar indicate the number and percentage of participants for each reason. Reasons included loss to follow-up (41%; 14 of 34), participant refusal or lack of interest in pursuing GT (26%; 9 of 34), awaiting consultation with a genetic counselor (24%; 8 of 34) or retinal specialist (3%; 1 of 34), lack of insurance coverage for testing (3%; 1 of 34), and other (3%; 1 of 34). The “other” category included one participant unable to complete home testing who cited GT as a low priority as they had no children to potentially inherit the condition.

### Factors associated with clinical trial (CT) participation in ABCA4-positive participants

Among 223 participants with positive or likely positive ABCA4*-*GT results, 20% enrolled in observational or interventional CTs, including TEASE^17^, ProgStar^18^, SMART^19^, SeaSTAR^20^, Polaris^21^, oral metformin^22^, Zimura^23^, DHA supplementation^24^, and Stargazer^25^. Multivariable analysis (**Figure 3**; complete numerical values available in **eTable 2**) showed later symptom onset was linked to greater CT participation; onset after age 60 had markedly higher odds (OR 36.662; 95% CI 2.502-537.143; p=0.009) compared to onset at 0-19 years.

**Figure 3.**
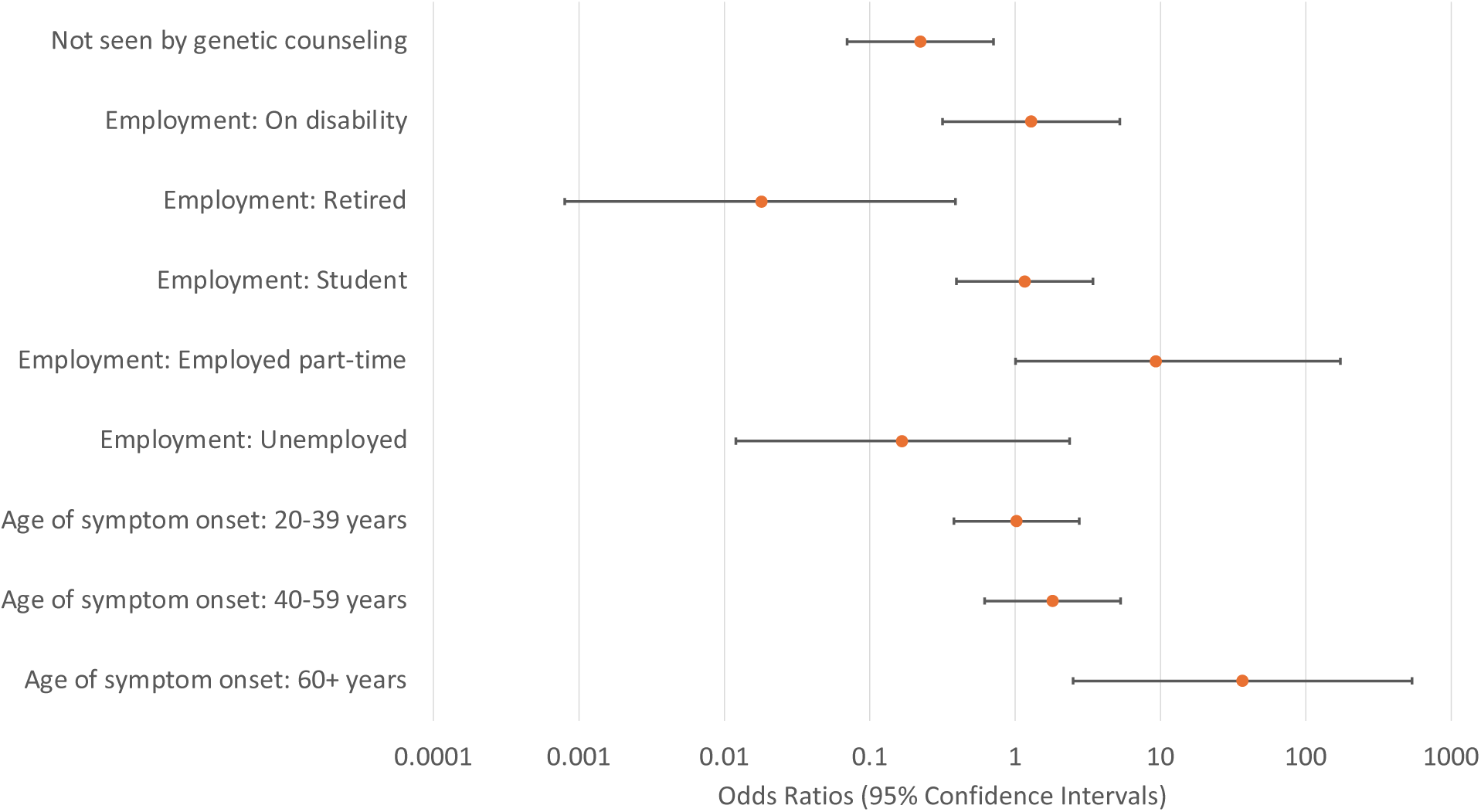
Multivariable analysis of factors associated with clinical trial participation among *ABCA4*-positive participants (N=223) Participants with confirmed positive (2 pathogenic variants, whether *in trans* or unknown phase) or likely positive (1 pathogenic + 1 VUS or 2 novel/VUS *in trans*, or 1 pathogenic variant [or two *in cis*] with typical Stargardt phenotype) *ABCA4*-specific genetic testing (ABCA4-GT) results were selected for further analysis to explore factors associated with clinical trial participation (n=223). Least absolute shrinkage and selection operator (LASSO) regression selected covariates for binary logistic regression. While there were 223 participants who had positive or likely positive genetic testing, the multivariable model includes 198 observations due to missing data in some variables. Odds ratios (ORs) and 95% confidence intervals (CIs) are plotted on a logarithmic scale, with a null value of OR = 1. Reference groups (REF) were selected by highest frequency: Age of symptom onset = 10-19 years; Employment = full-time employment.

Part-time employment was associated with greater odds of CT participation (OR 9.292; 95% CI 1.007-172.864; p=0.049), whereas retirement was associated with lower odds (OR 0.018; 95% CI 0.0008-0.389; p=0.011) compared to full-time employment. Lack of GC was again associated with reduced CT participation (OR 0.223; 95% CI 0.070-0.710; p=0.011).

## Discussion

Among participants with a molecular diagnosis of STGD at a genetic ophthalmology unit in a U.S. academic medical center, one-fifth had participated in a CT. CT participation was more likely among those with later symptom onset, part-time employment, and prior GC; conversely, retired individuals and those without GC were less likely to participate. Our findings provide a framework for principal investigators and study sponsors to understand potential barriers to completion cohort enrollment. Addressing these barriers systematically and rationally may ultimately help to increase CT participation among STGD subjects, thereby ensuring timely and complete study cohort enrollment.

We also found that 88% of STGD participants completed GT. Despite this encouraging rate, disparities persist. GT completion was less likely among Black participants, under/uninsured individuals, those with lower levels of educational attainment, those without plans for children, and those without prior GC. Although no-cost programs like eyeGENE and FFB-MRT-GTS were available to participants for the majority of the study period, lower completion rates among under/uninsured participants suggest that barriers extend beyond cost alone. Zhao et al. similarly found that while FFB-MRT-GTS improved GT access, uptake remained influenced by patient-level factors such as race and language preference, though no association was observed for insurance status.^9^ In contrast, studies in oncology have demonstrated that insurance status is a key determinant of GT access, with uninsured individuals significantly less likely to undergo testing.^26,27^ Together, these findings underscore the need to address both disease-specific and structural barriers to ensure equitable access to genetic services.

Individuals without children nor plans for children were also less likely to pursue GT, consistent with literature suggesting reproductive considerations are a major motivator for testing, with parents often more concerned about genetic risks to their children than to themselves.^28^ In contrast, those not planning to have children may perceive less relevance in testing. One untested participant in our cohort exemplified this view, stating they were uninterested in GT because they had no children to whom they could pass on their eye condition.

Bivariate analysis revealed that participants with post-secondary education (college, trade school, or graduate-level training) were significantly more likely to complete GT than those with only primary or secondary schooling. This aligns with prior research linking higher educational attainment to increased uptake of genetic services, likely due to factors such as enhanced health literacy and greater confidence in navigating healthcare systems.^29–31^

Common barriers to GT included delays in accessing GC, perceived cost, and limited interest or understanding of testing utility, particularly in the absence of FDA-approved treatments for STGD. Some participants initially expressed interest in GT but did not follow through, suggesting logistical or systemic obstacles such as delays in test ordering, limited follow-up, or challenges navigating the testing process. A single-center Australian study identified similar barriers to IRD testing, including delays in geneticist consultation and patient refusal.^11^

Black participants were significantly less likely to complete GT, reflecting known disparities in genetic testing uptake across specialties.^29^ Contributing factors may include reduced access to GC and GT, insurance coverage, provider referral patterns, and perceptions of GT shaped by historical and cultural experiences.^32^ Notably, lower diagnostic yield in Black patients has also been described in IRD literature, highlighting broader inequities throughout the GT process.^33^ Addressing these barriers requires expanding access to genetic services, diversifying genomic databases, and equipping providers with cultural competency training.^32,34^ Encouragingly, studies suggest that culturally sensitive communication approaches can enhance trust and engagement with genetic services among racially and ethnically diverse patients.^35^

Participants who received GC were significantly more likely to complete GT and enroll in CTs. CGCs may play a key role by informing patients of CTs and guiding them to speak with enrolling physicians—thereby facilitating GT uptake in partnership with the referring ophthalmologist, as is the practice at GEDi. Despite the clear benefit for CT participation, access to GC remains limited in many IRD care settings. A national assessment revealed high physician-to-CGC ratios and workforce shortages in ophthalmic GC, highlighting the need for expanded services.^36^ Additionally, although many ophthalmologists recognize the value of GT, barriers such as time constraints, incomplete knowledge of IRD genetics, and underutilization of GC referrals persist.^6^ Some eye care providers refrain from ordering GT due to the perception that it has little relevance for clinical management or treatment decisions.^6^ However, GT provides genotype-specific insights that inform CT (and future therapy) eligibility, family planning, and disease prognosis. Addressing this misconception and emphasizing GT’s broader benefits may increase GC referrals. To ensure equitable access to emerging therapies, it is essential to streamline referral pathways and incorporate GC more routinely into IRD care. Reducing diagnostic delays—which can span over a decade—and integrating GC early in the diagnostic journey can help patients make timely, informed, and supported decisions about GT and CT participation.^37^

Later symptom onset (≥60 years) and part-time employment were associated with higher CT participation, possibly reflecting greater time availability among part-time workers and increased interest in experimental treatments among participants with later-onset disease who may be more motivated by the potential for disease modification. Conversely, retired participants were less likely to participate, which may be attributable to concerns about the demands of CT involvement, competing health concerns, or lower perceived benefit given the progression of their disease. These findings suggest that factors such as employment status, age, and disease progression may influence patients’ willingness and ability to engage in clinical research. Understanding the unique factors that influence CT decision-making for eligible patients can help guide resource allocation and counseling strategies to enhance CT participation across diverse patient groups.

This study has several limitations. As a single-center retrospective study at a U.S. academic center, our findings may not generalize to other settings. There was considerable heterogeneity in participant follow-up, ranging from a single visit to longitudinal care spanning 21 years. CT participation may be underreported if participants enrolled in trials outside our institution or after transitioning care elsewhere. Additionally, not all participants were eligible for, or present at our institution during, the enrollment periods for each CT, and variable trial-specific criteria further limited comparability. Our aggregate, retrospective approach likely overestimates the number of truly eligible patients for any given CT. Due to variable documentation, we were unable to reliably determine for each CT how many patients were evaluated for eligibility, offered participation, declined enrollment, or were eligible but not offered the opportunity to participate. As a result, our CT participation estimates may not accurately reflect CT uptake among eligible patients. Finally, the retrospective design limited our ability to fully capture participant perspectives, including motivations for or against pursuing GT or CT enrollment.

As gene and cell-based therapies continue to advance, in tandem with pharmacological interventions for STGD, it will be crucial to implement rational strategies to promote CT participation. Future studies should incorporate prospective interviews or surveys to better understand systemic and individual barriers to GT and CT engagement. Moving forward, priorities should include expanding GC access, improving variant interpretation—especially in underrepresented populations—and fostering sustained engagement of patients and families in both clinical care and research.

## Conclusion

Our study demonstrates a high rate of GT completion among participants with STGD, as well as a 20% CT participation rate among those with positive or likely positive *ABCA4* results. However, disparities persist in GT completion—and, by extension, CT enrollment—particularly along racial and socioeconomic lines, highlighting critical gaps in equitable care. GT completion was significantly associated with race, insurance status, and clinical factors, underscoring the need for targeted interventions such as expanded no-cost GT initiatives and increased access to GC. Importantly, GC emerged as a key facilitator of both GT and CT participation, suggesting that integrating GC more consistently into the IRD care pathway could help mitigate disparities in GT access and support more equitable CT enrollment. Addressing these barriers is essential to ensuring that emerging IRD therapies are implemented equitably across all patient populations.

## Data Availability

All data produced in the present study are available upon reasonable request to the authors

## Financial Support

The Joseph Albert Hekimian Fund (MSS), Andreas C. Dracopoulos Professorship (MSS), Andreas C. Dracopoulos and Daniel Finkelstein M.D. Rising Professorship in Ophthalmology (JJD), Research to Prevent Blindness (unrestricted grant to the Wilmer Eye Institute)

## Data Access, Responsibility, and Analysis

DW and MSS had full access to all the data in the study and takes responsibility for the integrity of the data and the accuracy of the data analysis. Data analysis was conducted by DW (Johns Hopkins University School of Medicine).

## Data Sharing Statement

The data that support the findings of this study are available from the corresponding author, MSS., upon reasonable request.

## Conflict of Interest

No relevant financial disclosures.

## Notes

### Competing Interest Statement

The authors have declared no competing interest.

### Funding Statement

Financial support included The Joseph Albert Hekimian Fund (MSS), Andreas C. Dracopoulos Professorship (MSS), Andreas C. Dracopoulos and Daniel Finkelstein M.D. Rising Professorship in Ophthalmology (JJD), Research to Prevent Blindness (unrestricted grant to the Wilmer Eye Institute). The sponsor or funding organization had no role in the design or conduct of this research.

### Author Declarations

IRB of Johns Hopkins University gave ethical approval for this work

