## Supplemental Tables 1 and 2 for "Patient, Socioeconomic, and Clinical Influences on Clinical Trial Participation in Stargardt Disease"

### eTable 1. Multivariable logistic regression model of genetic testing completion

This table reports odds ratios (OR), 95% confidence intervals (CI), and exact p-values for covariates included in the model (corresponding to Figure 1). Predictors were selected via least absolute shrinkage and selection operator (LASSO) regression. Reference categories (REF) were selected based on highest observed frequency. The model includes all 280 participants. Null value = 1 for OR confidence intervals.

| Covariates | Odds Ratio (OR) | 95% Confidence Interval (CI) | P-value |
|---|---|---|---|
| **Race/ethnicity category** | | | |
| White | REF | – | – |
| Black | 0.395* | 0.160 – 0.976 | **0.044** |
| Asian | 0.961 | 0.169 – 5.457 | 0.964 |
| Hispanic or Latino | 1.133 | 0.106 – 12.137 | 0.918 |
| Other[a] | 0.871 | 0.079 – 9.566 | 0.910 |
| **Insurance status** | | | |
| Private insurance | REF | – | – |
| Under/Uninsured[b] | 0.468* | 0.193 – 0.972 | **0.043** |
| **Parenting status** | | | |
| Have or may plan to have children | REF | – | – |
| No children and no plans to have children | 0.348* | 0.137 – 0.885 | **0.027** |
| **Seen by genetic counseling** | REF | – | – |
| Yes | 0.085* | 0.033 – 0.221 | **4.18E-7** |
| No | | | |

[a]"Other" race was a composite category created for this analysis and includes Native Hawaiian or Other Pacific Islander and American Indian or Alaska Native, due to small sample sizes in each group. [b]"Under/Uninsured" status includes individuals with public insurance, no insurance, or international self-pay. *Statistically significant OR

### eTable 2. Multivariable logistic regression model of clinical trial participation among participants with positive or likely positive *ABCA4* results

Odds ratios (OR), 95% confidence intervals (CI), and exact p-values are reported for each predictor in the model (corresponding to Figure 3). Predictors were selected via least absolute shrinkage and selection operator (LASSO) regression. Reference categories (REF) were selected based on highest observed frequency. While there were 223 participants who had positive or likely positive genetic testing, the multivariable model includes 198 observations due to missing data in some variables. Null value = 1 for OR confidence intervals.

| Covariates | Odds Ratio (OR) | 95% Confidence Interval (CI) | P-value |
|---|---|---|---|
| **Age of symptom onset** | | | |
| 0-19 years | REF | – | – |
| 20-39 years | 1.024 | 0.380 – 2.763 | 0.962 |
| 40-59 years | 1.811 | 0.618 – 5.310 | 0.279 |
| 60+ years | 36.662* | 2.502– 537.143 | **0.009** |
| **Employment** | | | |
| Unemployed | 0.167 | 0.012 – 2.372 | 0.186 |
| Employed part-time | 9.292* | 1.007 – 172.864 | **0.049** |
| Employed full-time | REF | – | – |
| Student | 1.165 | 0.395 – 3.434 | 0.782 |
| Retired | 0.018* | 0.0008 – 0.389 | **0.011** |
| On disability | 1.290 | 0.317 – 5.244 | 0.722 |
| **Seen by genetic counseling** | | | |
| Yes | REF | – | – |
| No | 0.223* | 0.070 – 0.710 | **0.011** |

*Statistically significant OR
